# Enhancing the Positive Predictive Value of EGD for Diagnosis of Barrett’s Esophagus Through EsoGuard® Triage

**DOI:** 10.1101/2024.07.26.24311013

**Authors:** Jayde E. Kurland, Sheena B. Patel, Richard Englehardt, Seper Dezfoli, Daniel M. Tseng, Michael W. Foutz, Paul S. Bradley, Badi Eghterafi, Victoria T. Lee, Suman Verma, Brian J. deGuzman, Lishan Aklog

## Abstract

**Background:** Guidelines support Barrett’s esophagus (BE) screening, but most eligible patients do not undergo endoscopic evaluation; non-endoscopic strategies are now supported as a reasonable alternative by U.S gastroenterology societies. EsoGuard (EG) is a DNA assay used with EsoCheck, a non-endoscopic cell collection device for detection of BE, which can be utilized as a triage to esophagogastroduodenoscopy (EGD) in patients meeting screening criteria. In doing so, EG may serve to enrich the population undergoing EGD, resulting in more BE diagnoses while potentially reducing utilization of already-limited endoscopy resources.

**Aim:** To test the hypothesis that BE detection in EGDs performed on EG positive patients will be significantly higher than the positive predictive value (PPV) of screening EGD alone.

**Methods:** Real-world data was retrospectively collected from EG positive patients for whom EGD diagnoses were available. Baseline patient characteristics, risk factors, and EGD results were obtained from the treating physicians. PPV of screening EGDs was the comparator and estimated by literature-established disease prevalence of BE, which in the U.S gastroesophageal reflux disease population is ∼10.6%. The hypothesis was tested using t-tests for single proportions at a one-sided 5% significance level.

**Results:** Data from 209 patients found 60 (28.7%) subjects with salmon-colored mucosa on EGD and specialized intestinal metaplasia on histopathology. However, 10 (4.8%) had < 1cm of disease on visual inspection, therefore, did not meet the American College of Gastroenterology definition of BE so was excluded from the analysis. Of the remaining 199 patients, 50 (25.1%) had BE on EGD. In the cohort of patients meeting ACG screening criteria, 28.9% (33/114) had BE. Overall, a 2.4-fold increase in BE detection was observed compared to the PPV of screening EGD, and in the ACG cohort this increase was 2.7-fold. Among ACG patients ≥65 years old, the increase was nearly 2.5-fold (25.9% detection rate).

**Conclusions:** Our data suggests EG and EC used as a triage test enriches the population undergoing EGD for BE, and compared to screening EGD alone, can help direct more efficient use of endoscopy resources to unburden the system without reducing the number of eligible patients screened and diagnosed.

## Introduction

Barrett’s Esophagus (BE), a metaplastic condition of the lower esophagus, is the only known precursor for esophageal adenocarcinoma (EAC), the most common esophageal cancer in the U.S.[1] Among patients with BE, the risk of developing EAC is 30-152 times higher than in the general population.[2] In contrast to the lethality of EAC, which has an approximately 80% five-year mortality rate,[3] BE can be effectively treated with endoscopic eradication therapies (EET) such as radiofrequency or cryotherapy ablation which achieve complete eradication in up to 80-90%, underlining the need to screen and diagnose those at risk for BE.[4-7]

It is hypothesized that screening for BE, followed by surveillance to detect dysplasia (or early-stage carcinoma), and treatment of dysplastic BE with EET, may be the best strategy to reduce the incidence of EAC and EAC-related mortalities. As such, national societies like the American College of Gastroenterology (ACG) and American Gastroenterological Association (AGA) have published recommendations for screening patients at increased risk.[8, 9] The ACG endorses screening patients with chronic gastroesophageal reflux disease (GERD) and three or more of the following risk factors: male sex, White race, age >50 years, obese, history of tobacco smoking, and family history of BE/EAC in a first degree relative. BE is most frequently diagnosed when patients with refractory GERD undergo esophagogastroduodenoscopy (EGD); however, at-risk patients can be asymptomatic or have atypical symptoms.[10, 11] It should be noted that the use of acid suppressive medications is common; while these can reduce or even completely control GERD symptoms, the risk of BE is not entirely eliminated.[12, 13] These patients are unlikely to seek medical attention resulting in non-referral for endoscopic evaluation subsequently leading to missed BE diagnoses and missed opportunities to intervene before malignant progression. A study in U.S Veterans demonstrated that over 50% of patients with EAC did not report frequent GERD symptoms and would not have met ACG guidelines for BE screening.[14] As such, the AGA published recommendations in 2022 for screening patients with any three or more BE/EAC risk factors, without GERD as a prerequisite. BE/EAC risk factors such as male sex, White race, and age >50 years are highly prevalent and even GERD is seen in up to 44% of people in Western countries resulting in large numbers of at-risk patients.[15] Unfortunately, only about 10% of those appropriate for screening undergo EGD, because of obstacles that exist to performing screening EGDs on all eligible individuals.[16] Recognizing these limitations, both the ACG and AGA recommend non-endoscopic cell collection combined with DNA biomarker(s) testing as an acceptable alternative to EGD for screening of BE.[8, 9]

Non-invasive, in-office cell collection can allow wide-scale access to BE screening while also streamlining endoscopy resource utilization. In this workflow, the non-endoscopic cell collection combined with a DNA biomarker test serves as a triage step, and only those with a positive DNA test are referred for diagnostic EGD.[17, 18] EsoGuard (EG), a methylated DNA biomarker assay which analyzes esophageal cells collected with the FDA 510(k)-cleared swallowable EsoCheck (EC) balloon-capsule device, is the only such commercially available solution in the U.S.. Data from two National Cancer Institute (NCI)-funded case control studies demonstrated excellent sensitivity and specificity for detection of both BE and EAC using EG testing of esophageal cells collected with EC.[19, 20] More recently, a prospective, single-arm BE screening study at the Cleveland VA demonstrated EG sensitivity and negative predictive value (NPV) of 92.9% and 98.6% respectively on specimens collected with EC. The data also suggested that use of EG as an initial triage could increase the diagnostic yield of EGD by 2.5-fold.[21] To explore this in a non-veteran screening population, real-world data was collected retrospectively to test our hypothesis that the BE detection rate and therefore the positive predictive value (PPV) of EGD is increased among patients who first test positive with EG.

## Methods

### Study Design and Data Collection

The protocol for this retrospective study was submitted to the Institutional Review Board (IRB) and met requirements for a waiver of consent under 21 CFR 50.22. Data was collected from patients who underwent guideline-directed EG testing in the 2023 calendar year, whose test returned positive, and subsequent EGD results were also available. Patient demographics, BE/EAC risk factors, and final EGD diagnosis (plus biopsies where applicable) were accrued in a de-identified manner. Demographics and BE/EAC risk factors were collected from the test requisition forms (TRFs) stored in the database of the laboratory (Lucid Dx Labs, Lake Forest, CA) where the EG assay was performed. Older versions of the TRF only required age, sex, ICD-10 diagnosis codes, and documentation of the presence/absence of chronic GERD for processing, so for purposes of statistical analysis, missing or undocumented risk factors were treated as absent/negative. The TRFs also do not solicit duration of GERD symptoms, so it is assumed that patients for whom providers selected “chronic GERD” as a risk factor met ACG guideline definitions for chronicity (i.e., five or more years of symptoms).[8] Patients aged 65 years or older were categorized as “Medicare eligible.”

### EsoGuard and EsoCheck

EsoGuard (EG; Lucid Diagnostics Inc. New York, NY) is a targeted Next Generation Sequencing (NGS) DNA assay combined with a proprietary algorithm to examine presence of methylation on the vimentin (*VIM*) and Cyclin-A1 (*CCNA1*) genes. It has been analytically validated as a Laboratory Developed Test (LDT) performed in a Clinical Laboratory Improvement Amendments (CLIA) certified, College of American Pathologists (CAP) accredited, and New York State (NYS) Licensed laboratory (LucidDx Labs, Lake Forest, CA). EG utilizes a specific set of genetic assays and algorithms to examine the presence of cytosine methylation at 31 different genomic locations on *VIM* and *CCNA1*. Clinical validation of EG for detection of BE and EAC has been published from two NCI-funded case-control studies with pooled data demonstrating test sensitivity for EAC of 96% (95% CI: 80-99.9%), sensitivity for BE of 86% (95% CI: 76-90%), and specificity of 86 (95% CI: 81-91%).[19, 20]

EG is performed on cells collected non-endoscopically with EsoCheck (EC; Lucid Diagnostics Inc. New York, NY), an FDA 510(k)-cleared, swallowable capsule-balloon device designed for circumferential, targeted surface cell collection and protected retrieval of the specimen from the esophagus (**Figure 1**). Cell collection can be performed in any office setting without sedation and takes less than three minutes.[17, 20]

**Figure 1.**
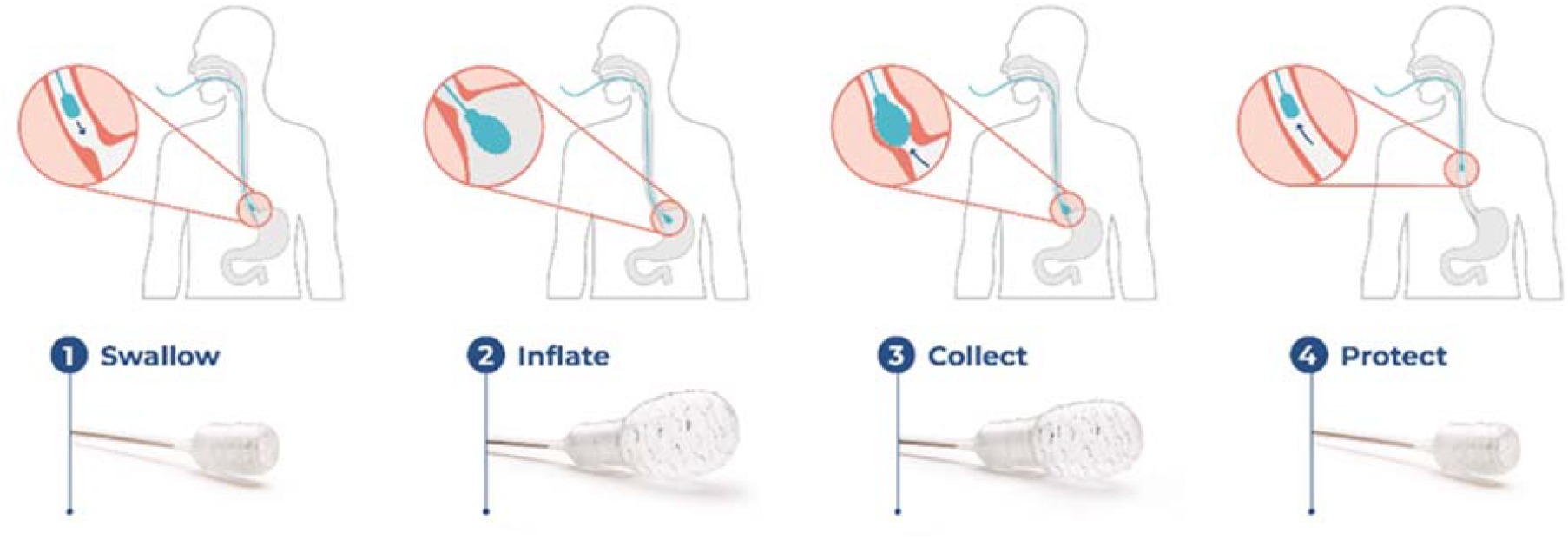
EsoCheck® Cell Collection Process.

### Classification of BE

The ACG defines BE as ≥1cm of columnar epithelium on EGD with histopathologic findings of specialized intestinal metaplasia (SIM).[8] In our study, patients with <1cm of non-dysplastic intestinal metaplasia on EGD, or non-dysplastic intestinal metaplasia confined to an irregular Z-line (otherwise known as specialized intestinal metaplasia of the esophagogastric junction; SIM-EGJ) were classified as “indefinite” for BE. Although SIM-EGJ fails to meet ACG length criteria for BE, the presence of SIM can appropriately trigger a positive EG, as biomarker tests are not specifically designed to distinguish disease length. Additionally, there is debate among experts about the clinical importance of detecting and endoscopically surveilling SIM-EGJ, as these lesions can and do progress to EAC, although risk of progression is lower for ultra-short segment disease.[22] In light of the above, patients diagnosed with SIM-EGJ were excluded from our primary outcome analysis. Patients with histopathologic findings of dysplasia were classified as positive for BE, irrespective of disease length.

### Statistical Analysis

Summary statistics were performed for demographics and other baseline characteristics. Measurement data were summarized using a mean with standard deviation and two-sided 90% confidence intervals (CI), and median with interquartile range (IQR). Chi-square was used for comparing categorical variables, and Kruskal-Wallace used for comparing group means. The primary aim/outcome of the study was to determine the disease detection rate (DR) of EGDs performed in EG-positive patients, and to assess whether this is superior to the PPV of screening EGDs alone. DR represents the PPV of EGDs performed *after a positive EG result*, while the PPV of screening EGDs is expected to be equal to the BE prevalence in the overall at-risk population. A meta-analysis of the U.S. GERD population showed this prevalence to be 10.6%, which serves as the performance goal of our primary outcome analysis.[23] Prevalence range is cited as 5-15% in other literature making 10.6% a reasonable and conservative estimate.[1]

We hypothesize that triaging patients with EG prior to EGD will significantly increase the DR of those EGDs beyond the 10.6% performance goal. The study hypothesis was tested using t-tests for single proportions at a one-sided 5% significance level. The outcomes for the primary study aim are presented using point estimates for frequency and percentages and normal two-sided 90% CI.

## Results

Among approximately 10,000 patients who underwent EsoGuard in 2023, data was successfully collected from 209 of the patients who had both positive test results and for whom the ordering providers willingly shared de-identified findings from the subsequent EGDs. There were 51 unique ordering providers, and the EGDs were performed by 78 different endoscopists. Thirteen of the ordering providers (25.4%) were the endoscopists for their own EG positive patients, while the remainder were primary care providers (**Figure 2**). Geographically, 44.0% (92/209) of the tested patients were from the Eastern U.S (states east of Mississippi); 35.8% (75/209) were from the Western U.S (states west of Colorado), and 20.2% (42/209) from the Central U.S (any states in between).

**Figure 2.**
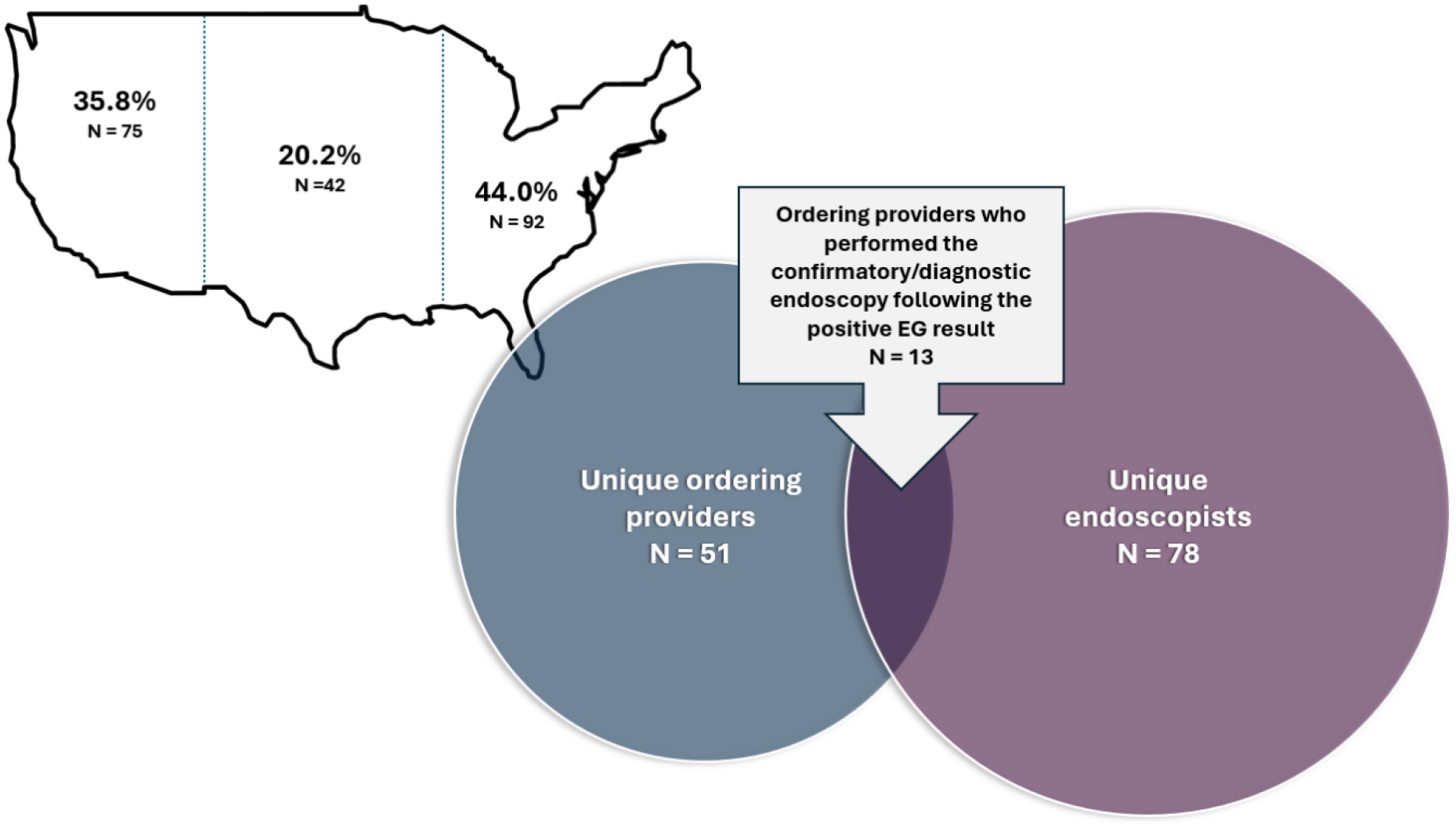
Geographic Distribution and Characteristics of EsoGuard Ordering Providers Contributing Data to the Study.

Patient baseline characteristics are summarized in **Table 1**. The average age was 64.1 years, and not significantly different among BE positive vs. BE negative patients. Over 54% (113/209) were 65 years or older, contributing to the cohort of “Medicare-eligible” patients. Male sex was more prevalent among the BE positive cohort (72.0%) than the BE negative (54.4%), but this did not meet statistical significance. The proportion of chronic GERD patients was slightly higher among the BE negative cohort (81.9%) than the BE positive (74.0%). The most common risk factors were age ≥50 years (88.0%), White race (83.3%), and chronic GERD (78.5%). Approximately 50% percent of patients were obese, 56.1% had a tobacco smoking history, and 8.9% had a family history of BE or EAC in a first degree relative. There was a statistically significant increased prevalence of tobacco smoking among BE positive (72.3%) and SIM-EGJ patients (77.8%), compared to BE negative (49.3%). Although not included as a risk factor in the ACG or AGA screening criteria, 42 (20%) of the tested patients were firefighters, an occupation with known significant increased risk of developing EAC.[24, 25] These individuals were likely tested as part of department-led screening events. Firefighter representation did not significantly differ between the BE positive vs. BE negative groups.

**Table 1.**
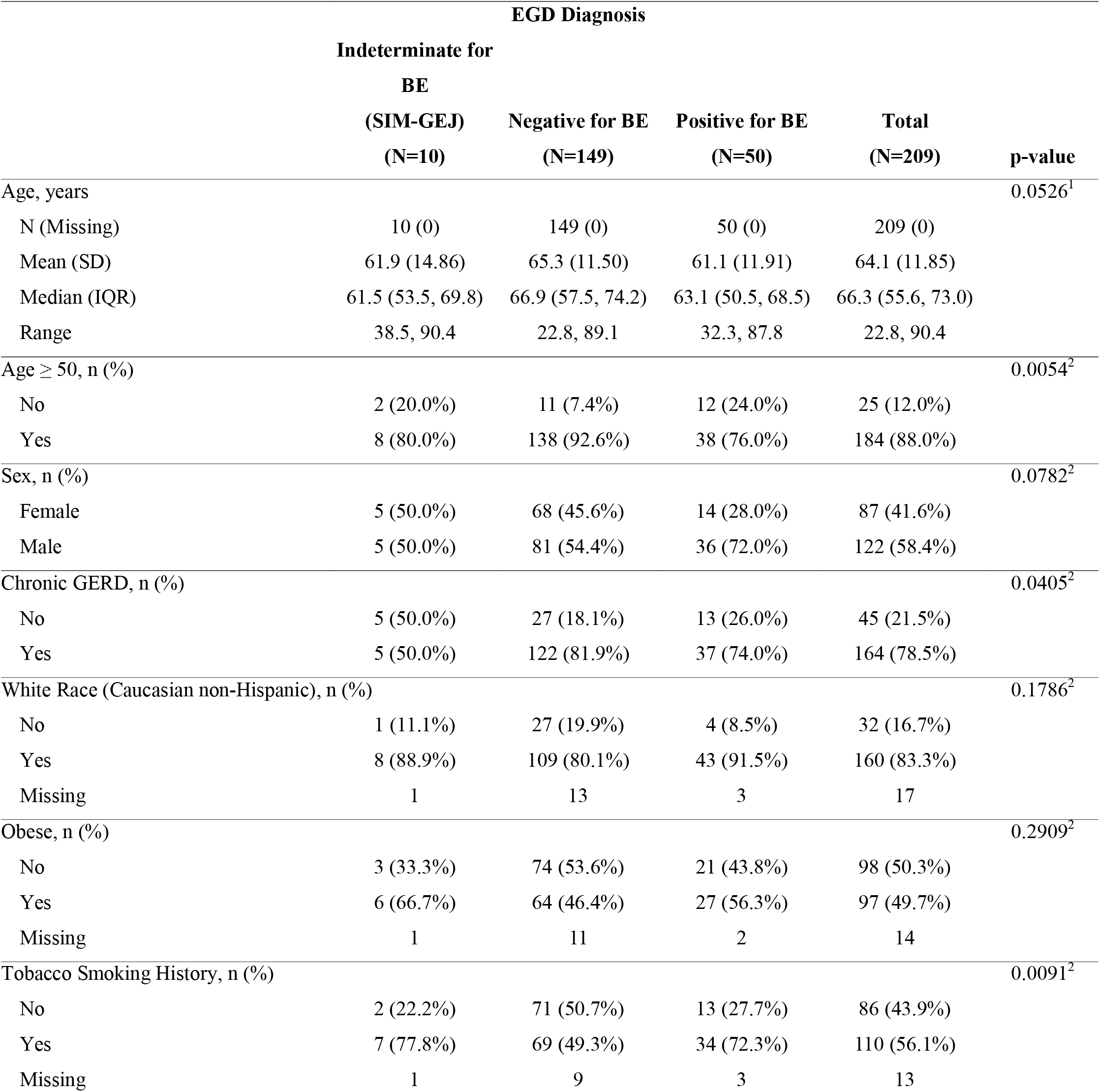
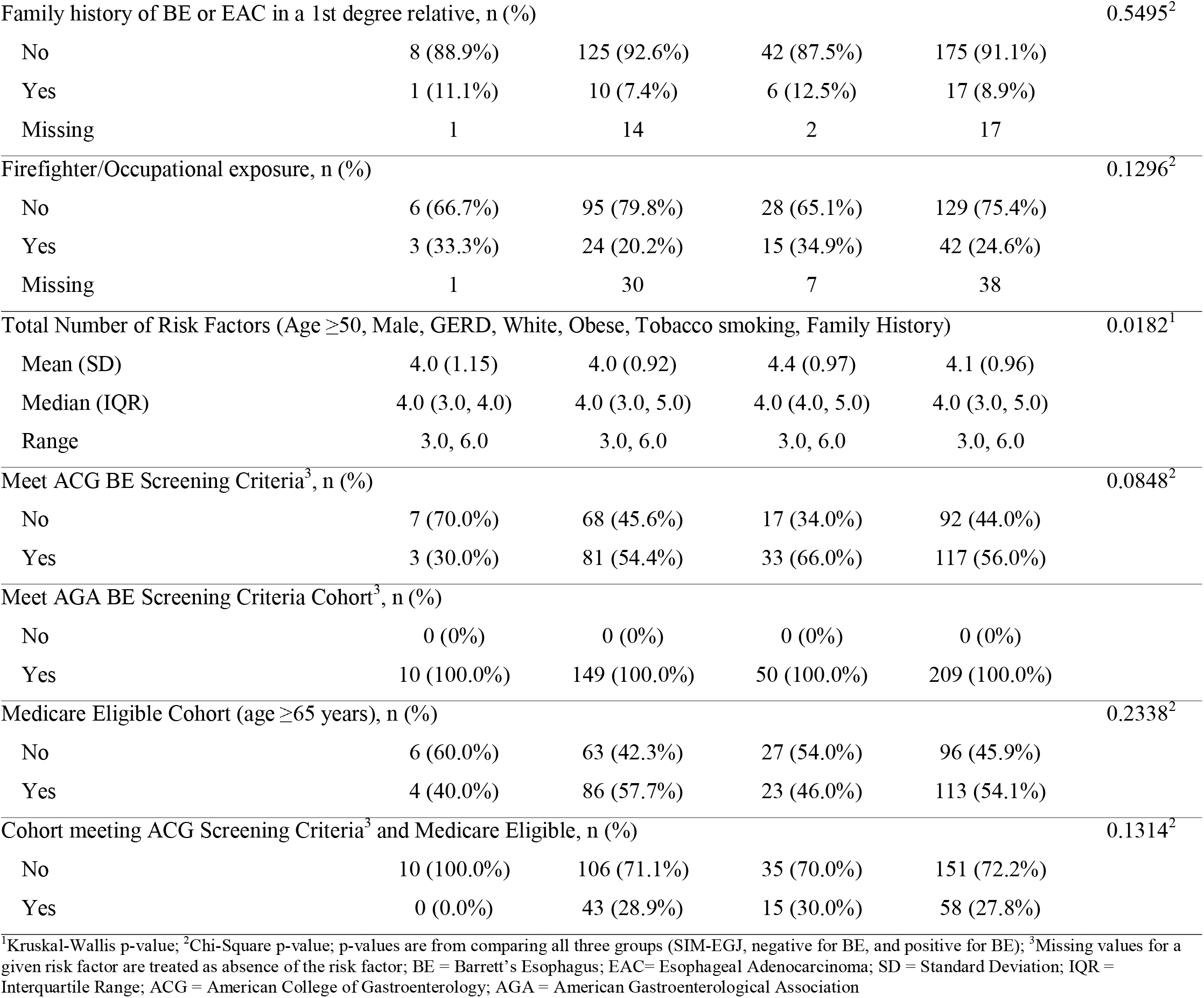
Demographics and Baseline Characteristics by EGD Diagnosis: comparison of SIM-EGJ, Negative for BE, and Positive for BE.

The average number of risk factors was higher in patients with BE than those without, and as expected, this group also had a higher proportion of patients meeting ACG guideline criteria for screening (66.0%, compared to 54.4% in the BE negative group) although the latter did not meet statistical significance. In the study 56% of subjects met both ACG and AGA screening criteria, with the remaining 44% meeting AGA criteria without reaching the threshold for ACG.

In **Table 2** the EGD results from patients meeting ACG BE screening criteria are compared to results of patients meeting *only* AGA criteria, with EGD diagnoses of the overall study population provided for reference. Of the EG positive patients, BE was detected in 23.9% (50/209) and not detected in 71.3% (149/209); 4.8% (10/209) had SIM-EGJ and classified as “indeterminate” for BE. Patients meeting ACG screening criteria were compared to those meeting AGA but not ACG criteria; the ACG cohort had a higher rate of BE on EGD (28.2% vs. 18.5% respectively) and lower rate of SIM-EGJ (2.6% vs. 7.6% respectively), although this difference was not statistically significant. Distribution of disease stage did not significantly differ between the cohorts. Most disease was non-dysplastic BE (NDBE; 84.0%, 42/50; full study population), and there were no cases of cancer.

**Table 2.** Disease Detection on EGD in EsoGuard Positive Patients: Comparison of Patients Meeting ACG Screening Criteria vs. Patients Meeting only AGA Screening Criteria.

|  | Overall (N= 209)*<br>[90% CI] | Patients meeting ACG<br>Guideline Criteria for BE<br>Screening<br>(n=117)<br>[90% CI] | Patients meeting only AGA<br>Guideline Criteria for BE<br>Screening§<br>(n=92)<br>[90% CI] | p-value |
| --- | --- | --- | --- | --- |
| **Diagnosis category (positive for BE, indeterminate for BE, negative for BE)** |  |  |  | 0.085<sup>1</sup> |
| Indeterminate (SIM-EGJ) | 4.8% (10/209)<br>[2.6%,8.0%] | 2.6% (3/117)<br>[0.7%,6.5%] | 7.6% (7/92)<br>[3.6%,13.8%] |  |
| Negative | 71.3% (149/209)<br>[65.7%,76.4%] | 69.2% (81/117)<br>[61.5%,76.2%] | 73.9% (68/92)<br>[65.3%,81.3%] |  |
| Positive | 23.9% (50/209)<br>[19.1%,29.3%] | 28.2% (33/117)<br>[21.4%,35.8%] | 18.5% (17/92)<br>[12.1%,26.4%] |  |
| **Stage of disease on EGD and biopsies** |  |  |  | 0.609<sup>1</sup> |
| Non-dysplastic BE<br>(NDBE) | 84.0% (42/50)<br>[73.0%,91.8%] | 81.8% (27/33)<br>[67.2%,91.8%] | 88.2% (15/17)<br>[67.4%,97.9%] |  |
| Indefinite for dysplasia | 6.0% (3/50)<br>[1.7%,14.8%] | 9.1% (3/33)<br>[2.5%,21.9%] | 0.0% (0/17)<br>[0.0%,16.2%] |  |
| BE with low-grade<br>dysplasia (LGD) | 4.0% (2/50)<br>[0.7%,12.1%] | 3.0% (1/33)<br>[0.2%,13.6%] | 5.9% (1/17)<br>[0.3%,25.0%] |  |
| BE with high-grade<br>dysplasia (HGD) | 6.0% (3/50)<br>[1.7%,14.8%] | 6.1% (2/33)<br>[1.1%,17.9%] | 5.9% (1/17)<br>[0.3%,25.0%] |  |

<sup>1</sup>Chi-Square p-value, p-values are from comparison of the ACG to AGA non-ACG groups; CI = Confidence Interval; SIM-EGJ = Specialized Intestinal Metaplasia of the Esophagogastric Junction; AGA = American Gastroenterological Association; ACG = American College of Gastroenterology; BE= Barrett’s Esophagus; NDBE = Non-dysplastic Barrett’s esophagus; LGD = Low-grade dysplasia; HGD = High-grade dysplasia
§By definition, all patients meeting ACG criteria also meet AGA criteria for BE screening

When patients with diagnosis of SIM-EGJ are excluded from the primary outcome analysis, the evaluable N decreases to 199. Among these, 25.1% (50/199) were positive for BE. The DR for different cohorts is summarized in the forest plot of **Figure 2**. DR of EGD in patients who triaged to EGD with a positive EG result was higher than the performance goal of 10.6% (dotted vertical line, **Figure 2**) in all cohorts, but was highest in those who met ACG screening criteria (28.9%). In the subset of patients meeting ACG BE screening criteria who were also Medicare eligible (i.e., aged ≥ 65 years), DR was 25.9%. A flow chart of patient allocation into the different cohorts and the respective primary outcomes (DR) is provided in **Supplemental Figure 1**.

**Figure 2.**
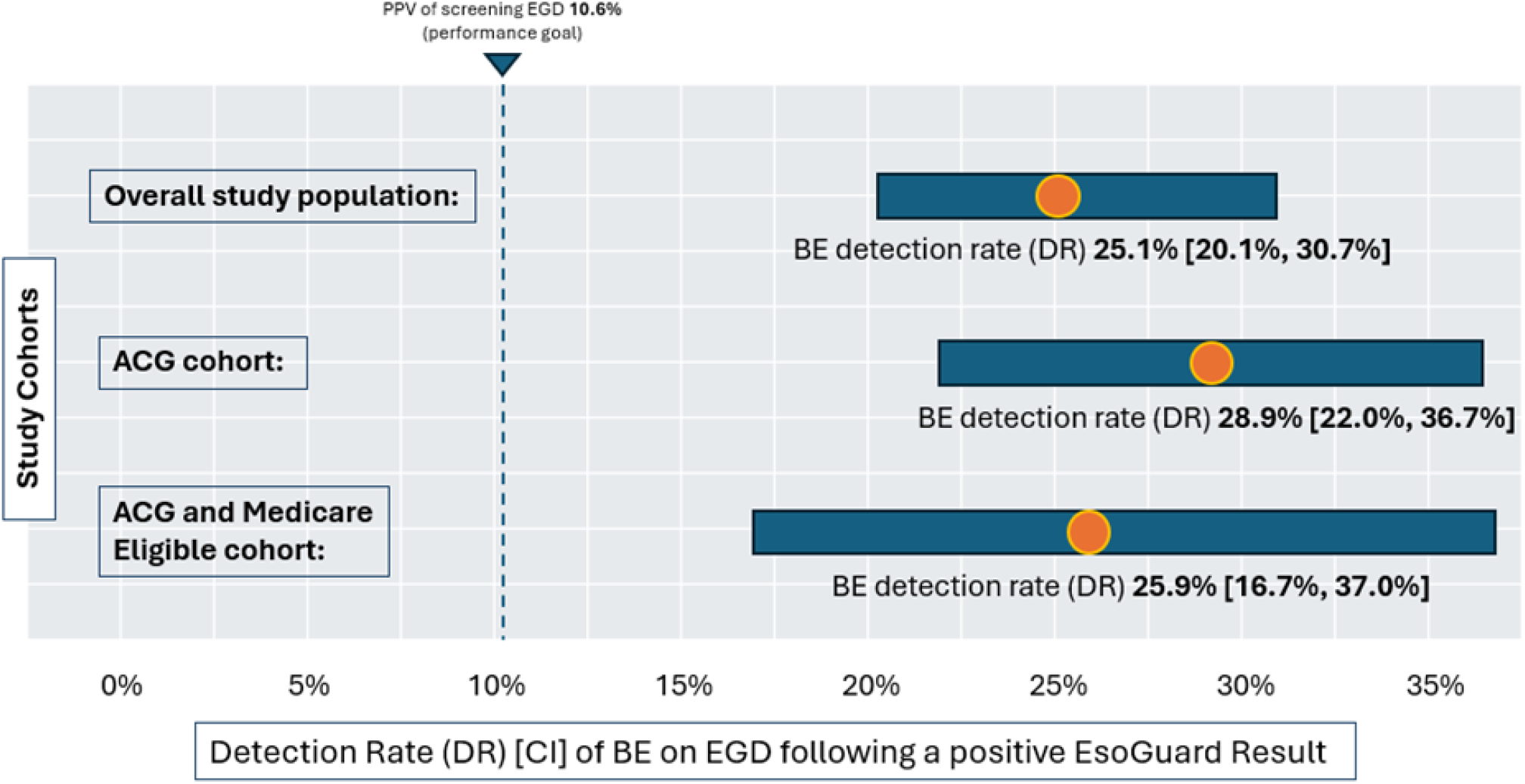
Positive yield of EGD after EG positive result in major population cohorts of the study. ACG = American College of Gastroenterology; AGA = American Gastroenterological Association; DR = Detection Rate The positive predictive value (PPV) of EGD performed in the EG positive population is represented for each cohort by disease detection rate (DR). DR represents the PPV of EGDs performed after a positive EG result. The dotted line denotes the 10.6% performance goal for all study cohorts, representing the estimated PPV of screening EGD which is based on disease prevalence in the U.S GERD population;[23] the Medicare Eligible cohort included patients aged 65 years or older.

The DR of EGD in EG positive patients was also calculated among cohorts with specific risk factors (**Figure 3**), namely: obesity, male sex, GERD, family history of BE/EAC in a first degree relative, and age 50 years or older. These were compared to the PPV of screening EGD, which was based on literature-established BE prevalence for each risk cohort[26]. In all risk cohorts, the DR of EGD in EG positive patients was significantly higher than the primary performance goal of 10.6%. In all risk cohorts except those with a family history, the DR was significantly higher than the prevalence of disease/PPV of screening EGD.

**Figure 3.**
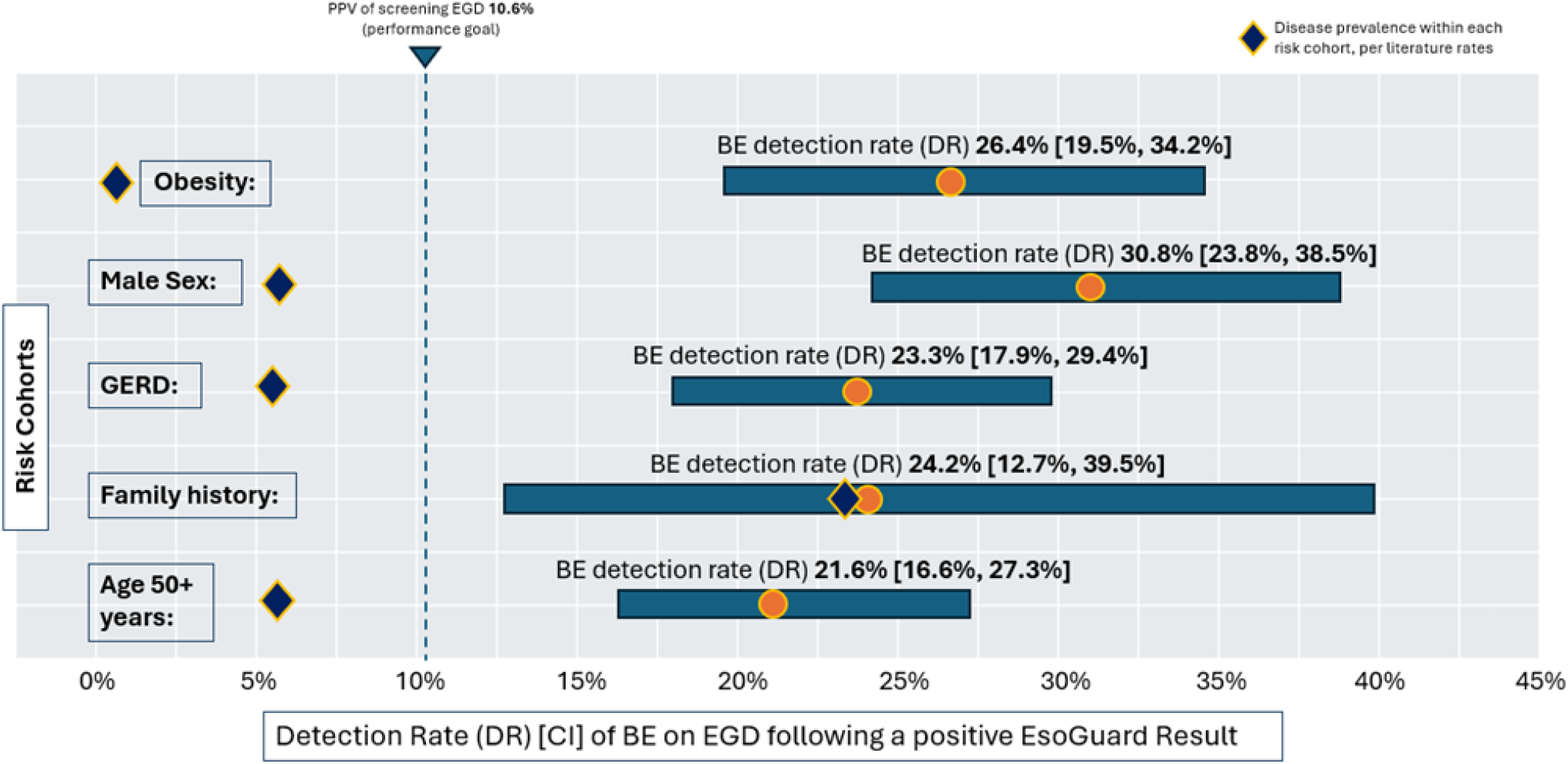
Positive yield of EGD after EG positive result in cohorts with specific risk factors. GERD = Gastroesophageal Reflux Disease; DR = Detection Rate The positive predictive value (PPV) of EGD performed in the EG positive population is represented for each cohort by disease detection rate (DR). DR represents the PPV of EGDs performed after a positive EG result. The blue diamonds denote disease prevalence for each risk population based on published meta-analysis data, representing the PPV of a screening EGD alone. Prevalences are as follows: Obese, 1.9%; Male sex, 6.8%; chronic GERD, 6.2%; family history of BE or EAC in a first degree relative, 23.4%; Age 50 years or older, 6.1%. [26] The dotted line denotes the study’s 10.6% DR performance goal.

## Discussion

This study presents real-world data from patients who underwent guideline-supported BE screening with non-endoscopic cell collection using EsoCheck and the methylated DNA biomarker assay, EsoGuard, followed by EGD in those who tested positive. The detection rate (DR) of BE in these patients represents the PPV of EGDs performed *after EG triage*, which we compared to the PPV of screening EGD alone.

These data support our hypothesis that the PPV of EGD for detecting BE can be significantly improved by triaging patients with the combination of EG and EC. Our findings are consistent with a recent study by Greer et. al. where disease prevalence in the Cleveland VA screening population meeting ACG criteria was 12.9%, and the PPV of EGD was increased 2.5-fold in those who first triaged positive with EG.[21] These data suggest that EG, when used as a first step in BE screening, can enrich the population undergoing EGD and in doing so, direct more efficient utilization of endoscopy resources.

BE was detected on EGD in 25.1% of our study population, and 28.9% in the cohort specifically meeting ACG screening criteria. This is a 2.4-fold and 2.7-fold increase above the performance goal, respectively. The study’s performance goals assumed the PPV of screening EGDs (performed *without* biomarker triage) will match the population’s disease prevalence, which in U.S GERD patients is about 10.6%.[23] This estimate sits well within the 5-15% prevalence range published in the literature for the overall at-risk population.[1] However, while a population’s BE prevalence is a good proxy for the PPV of screening EGD, in reality only ∼10% of individuals eligible for screening ever undergo endoscopic evaluation.[16] Low rates of primary care referrals for screening EGD, patient access and logistical barriers to scheduling EGDs, and patient fear of the procedure are all contributory.[27, 28] Thus, although EG triage may increase PPV of EGD by 2.4 to 2.7-fold, the impact on increased disease detection could be as high as 22 to 24-fold, based on the fact that in contrast to the 10% of screening-eligible patients who undergo EGD as the first-line diagnostic test, we estimate up to 90% of patients would be willing to undergo EG. This is supported by survey data from the technology’s pivotal study, where >90% of surveyed patients reported a willingness to repeat EG/EC testing, and >90% stated they would recommend this testing to others.[19] Additionally, published data from an ongoing EsoGuard registry demonstrates that patients with positive test results are reliably being referred for the recommended confirmatory EGD, thus minimizing missed diagnoses at this step.[17] By substantially improving accessibility and compliance with screening, EsoGuard can exponentially increase current rates of BE detection.

We observed that the subset of Medicare eligible patients within the ACG cohort had a slightly lower DR at 25.9% (vs. 28.9%), potentially suggesting that EG performance in elderly patients may differ from those who are younger. This finding may be attributed to potentially lower EG specificity in older patients, as was seen in the Barrett’s Esophagus Translational Research Network (BETRNet) study, where patients aged >61 years had test specificity of 76% whereas younger patients had test specificity of 81%.[20] This is hypothesized to be due to increased epigenetic changes including alterations in DNA methylation that occur with advanced age. While aging in mammals is more commonly associated with hypomethylation, DNA methylation can also increase over the CpG islands of certain genes, in a nonstochastic manner.[29] Additionally, a study of colonic mucosa suggested that aging could be a major contributing factor to the hypermethylation seen in cancer.[30] Aberrant methylation of the *VIM* and *CCNA1* genes are known to be associated with gastrointestinal pathologies, but it is unknown whether they are among the genes which experience age-related hypermethylation; such a phenomenon would lead to lower specificity of any methylation-based biomarker assays performed in the elderly population.[31] [19] However, since the DR in the Medicare-eligible subset of ACG patients is 2.5 times higher than the performance goal, it indicates that EG triage is still effective at improving the diagnostic yield of endoscopy in older individuals. Compared to screening EGD alone, the “number needed to test” with EsoGuard to detect each additional case of BE would be 6.5 in the Medicare-eligible ACG cohort, and 5.5 in the full ACG cohort.

We evaluated the DR of cohorts with specific BE/EAC risk factors and compared this to the PPV of screening EGD (modified according to published disease prevalence in each risk cohort).[26] In the obese population, males, population with chronic GERD, and those aged ≥50 years, the DR of EGD in EG positive patients was significantly higher than the PPV of screening EGD alone. However, the impact of positive EG results on improved PPV is difficult to quantify, as cohorts were not analyzed in a mutually exclusive fashion and the presence of additional risk factors within each cohort was pervasive.

As expected, chronic GERD was a highly prevalent risk factor among our study population, given that all patients met either ACG or AGA risk criteria for BE screening. Interestingly however, the GERD risk factor was not proportionally higher in the study cohort with BE on EGD as compared to those who were BE negative. Chronic GERD is defined by both the ACG and AGA guidelines as ≥ 5 years of symptoms that occur at least weekly, although the two societies have differing positions on whether it should be a prerequisite for BE screening. The AGA recognized in their guidelines that a meaningful proportion of patients who develop BE and EAC do not report heartburn or other reflux symptoms, nor carry a diagnosis of chronic GERD. This is consistent with what we observed here as 21.5% (45/209) of patients diagnosed with BE on EGD did not have GERD documented as a risk factor. Indeed, a study of EAC in patients from the U.S and U.K demonstrated that 54.9% of the U.S EAC patients would not have met ACG guideline criteria for screening, 86.5% of which would have been due to the absence of symptomatic GERD.[32] Our data suggest that patients meeting either ACG or AGA guideline criteria for BE screening could benefit from non-endoscopic biomarker testing strategies like EG/EC.

The vast majority of BE cases in our study were NDBE (84%), consistent with patterns seen in the literature where NDBE accounts for anywhere from 70 to >90% of the BE population.[23, 33]Indefinite for dysplasia (IFD) and LGD accounted for 10% and HGD accounted for 6% of our cases, which aligns with the 11.5% LGD, and 5.1% HGD for 5.1% seen in a publication from a large multi-center consortium.[33] While no EAC cases were represented in our study population, this is also expected based on the low population prevalence, which is reported at 0.6% in U.S GERD patients.[23] This suggests our study population is a good representation of the real-world BE population, and our findings are unlikely to have been significantly impacted by bias from the retrospective nature of data collection and analysis.

Our study has several limitations. First, BE/EAC risk factors were incompletely collected for 17 patients, since information was obtained retrospectively from TRFs. Although documentation of risk factors was an option in older versions of the TRF, it was not until April of 2023 that it became required for assay processing. Prior to this time, only age, sex, and ICD-10 diagnosis code (used to identify patients with chronic GERD) were mandated. We took a conservative approach when categorizing patients into the ACG and AGA cohorts, and unless a risk factor was specifically documented as being present, it was otherwise inferred as being absent or negative; as such, the number of patients meeting ACG guideline criteria for BE screening are likely underestimated in our analysis. However, given the consistency of our findings with those of Greer et. al., whose patient population also met ACG screening criteria, we believe any impact on the primary outcomes of our study would not be substantial.

A second limitation is the fact that our study included only a small subset of EG positive patients for whom we were able to obtain EGD results. The possibility of sampling bias cannot be entirely eliminated, although the geographic distribution of participants and ordering providers suggests the data arises from a random subset of the commercial patients with positive EG results. The regional representation seen in our study’s population is a reasonable reflection of the broader U.S population distribution, with higher numbers along either coast, and slightly lower numbers in the central portion of the country. Finally, although we hypothesize that in-office testing options like EG and EC should increase screening by facilitating patient access and potentially addressing health equity issues associated with EGD, future studies are necessary to quantify this impact, particularly in rural and under-served areas.

## Conclusion

In summary, among patients who first tested positive with EsoGuard, the BE detection rate on subsequent EGDs was 2.4 to 2.7-fold higher than the PPV of screening EGD alone in an at-risk population. The increased detection rate was greatest in the cohort meeting ACG guideline criteria for screening. Within the subset of this cohort meeting Medicare age eligibility, the detection rate was still approximately 2.5-fold higher than the PPV of screening EGD, despite potential of age-related aberrant DNA methylation to impact specificity in methylation-based biomarker assays. This demonstrates that non-endoscopic cell collection and triage with EG effectively enriches the population undergoing EGD for diagnosis of BE. Compared to EGD alone as the sole screening tool, the EG biomarker-based triage test can help direct more efficient use of endoscopy resources and unburden the system without reducing the number of eligible patients being effectively screened and diagnosed.

## Data Availability

All data produced in the present study are available upon reasonable request to the authors

