## Supplemental Figure 1 for "Enhancing the Positive Predictive Value of EGD for Diagnosis of Barrett’s Esophagus Through EsoGuard® Triage"

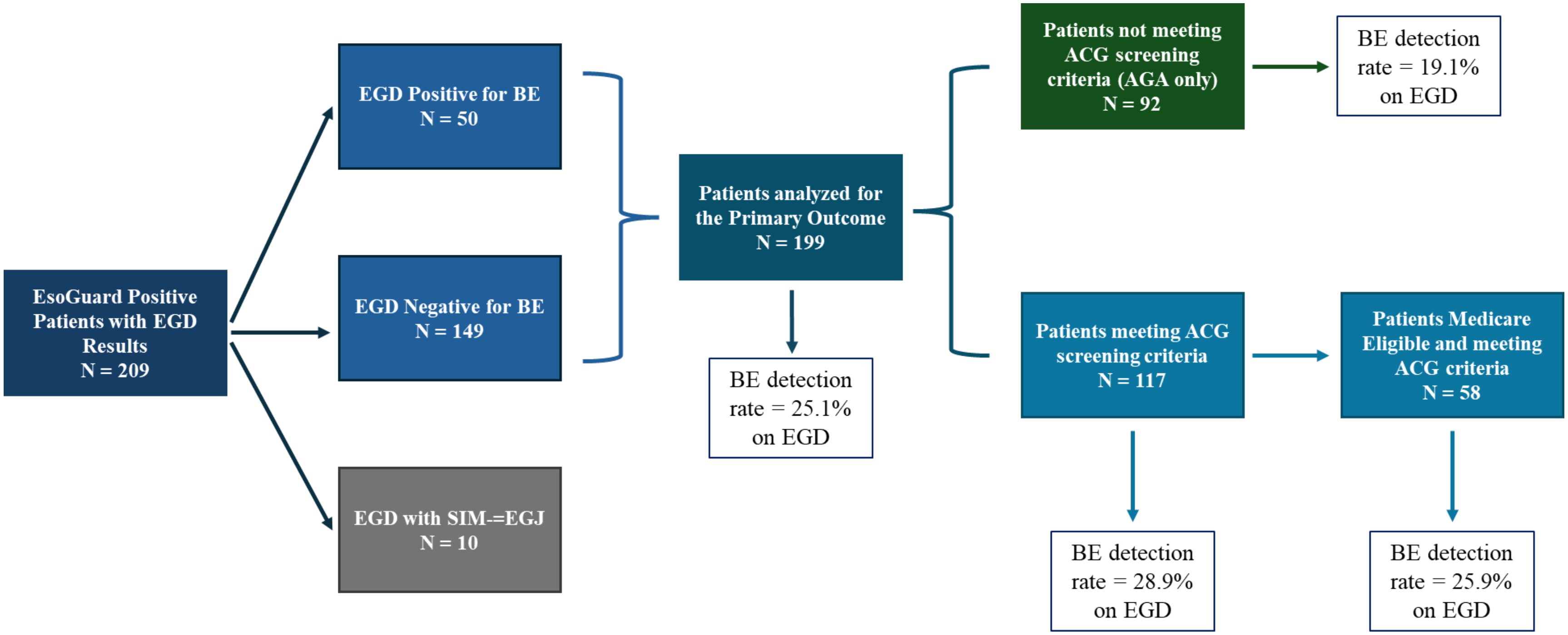


**Supplemental Figure 1.** Patient flow chart and associated primary outcomes

EGD = esophagogastroduodenoscopy; BE = Barrett's esophagus; SIM-EGJ = Specialized Intestinal Metaplasia of the Esophagogastric Junction; AGA = American Gastroenterological Association; ACG = American College of Gastroenterology
